# Machine Learning Enabled Non-invasive Diagnosis of Nonalcoholic Fatty Liver Disease and Assessment of Abdominal Fat from MRI Data

**DOI:** 10.1101/2022.03.25.22272965

**Authors:** Arvind Pillai, Kamen Bliznashki, Emmette Hutchison, Chanchal Kumar, Benjamin Challis, Mishal Patel

## Abstract

Nonalcoholic fatty liver disease (NAFLD) is the most rapidly growing contributor to chronic liver disease worldwide with high disease burden and suffers from limitations in diagnosis. Inspired by recent advances in machine learning digital diagnostics, we explored the efficacy of training a neural network to classify high risk NAFLD vs. non-NAFLD patients in the UK Biobank dataset based on proton density fat fraction (PDFF). We compared the performance of several ResNet-derived architectures in the context of whole abdomen MRI, segmented liver and abdomen excluding liver (sans-liver). Non-local ResNet trained on whole abdomen MRI images yielded the highest precision (0.88 for NAFLD) and F1 (0.89 for NAFLD). Furthermore, our work on a second, larger cohort explored multi-task learning and the relationship among PDFF, visceral adipose tissue (VAT) and abdominal subcutaneous adipose tissue (ASAT). Interestingly, multi-task learning experiments found a decline in performance for PDFF when combined with VAT and ASAT. We address this deterioration using Multi-gate Mixture-of-Experts (MMoE) approaches. Our work opens the possibility for using a non-invasive deep learning-based diagnostic for NAFLD, and directly enables clinical and genomic research using a larger cohort of potential NAFLD patients in the UK Biobank study.

## I. Introduction

Nonalcoholic fatty liver disease (NAFLD) is a hepatic manifestation of metabolic disorders characterized by excess accumulation of fat in hepatocytes [1]. Recent estimates suggest that NAFLD may be affecting more than 25% of global population and there is considerable variability in the prevalence of NAFLD across the various geographic regions in the world [2]. A subset of NAFLD patients develop advanced forms of liver disease such as nonalcoholic steatohepatitis (NASH), and fibrosis, which can potentially progress to cirrhosis [1], [3]. The current standard to diagnose NAFLD is the liver biopsy, a costly and invasive procedure with associated morbidity, poor patient tolerability and sampling variability [4].

As a result, liver biopsy is not practical for screening large populations of at-risk individuals, or for monitoring changes in fibrosis stage over time or in response to novel therapies [5]. There is an unmet medical need to develop non-invasive and precise biomarkers for objective and accurate disease diagnosis, and patient stratification.

Magnetic resonance imaging (MRI) now provides a promising non-invasive diagnostic alternative that can be used for large scale population studies and for serial follow up of patients at risk. MRI techniques allow comprehensive and objective evaluation of NAFLD [6]. To non-invasively assess NAFLD, Lin *et al*. used radiofrequency ultrasound data to classify patients with proton density fat fraction (PDFF) greater than 5% [7]. MRI-PDFF has been demonstrated to be a reliable method to clinically estimate liver fat [8], [9]. In addition to PDFF, visceral adipose tissue volume (VAT) and abdominal subcutaneous adipose tissue volume (ASAT) provides useful information for assessing NAFLD. A recent study by Jung *et al*. shows high visceral to subcutaneous fat ratio is associated with increased NAFLD risk [10].

Deep learning has been used to address central problems in medical imaging [11]. In abdominal imaging, convolutional neural networks (CNNs) like the U-Net have been used to segment liver, kidney, spleen, and pancreas effectively [12]-[14]. Extracting useful information for liver disease diagnostics has also gained traction. For instance, Yasaka *et al*. used CNNs to successfully predict five different fibrosis stages from phase MRI [15].

Combining PDFF, VAT, and ASAT features to identify NAFLD is a promising direction for research. Predicting these features as separate tasks from the abdomen MRI data using distinct models, however, is neither practical nor computationally efficient. Multi-task learning (MTL) offers a powerful framework to simultaneously predict several tasks.

Using this paradigm for diagnostic medical imaging has provided significant improvements in performance over standalone models [16]-[18]. In cases where the relationship between tasks is complex it can be difficult to assess if there is a positive transfer between tasks, i.e. adding an extra task and training a MTL model improves performance over the standalone model.

To address these questions, we first developed a 3D CNN using multi-echo spoiled-gradient-echo MRI from 4,607 subjects in the UK biobank to accurately (F1 = 0.89) classify subjects with NAFLD (PDFF > 5.5%). Following this we performed architecture search, benchmarking state of the art 3D classification models. The contribution of the liver towards accurate classification of NAFLD using PDFF was examined first by developing and validating a segmentation model to segment the liver from the abdomen followed by performing the same classification on abdomen, liver-only and abdomen excluding liver (referred as sans-liver) data. Based on these findings we then examined the contribution of auxiliary variables VAT and ASAT to assess the possibility of improvement in prediction of PDFF and classification of NAFLD. We developed a multitask learning, multi-gate mixture of experts 3D CNN using a larger cohort of 9,814 subjects from the UK biobank to predict raw values of PDFF, VAT, and ASAT. We compare the multitask approach with standalone models for each variable and measure task similarity and learning transfer among the prediction tasks. Finally, we analyze the importance of the liver and connection to the auxiliary variables on the segmentation data.

## II. Methods

### A. UK Biobank Liver MRI Data

In this study, we use multi-echo spoiled-gradient-echo abdominal MRI data from the UK Biobank [19]. Each image volume has 160 × 160 acquisition matrix with 10 slices (depth), with pixel dimensions and slice thickness of 2.5 × 2.5 *mm* and 6 *mm*, respectively. We used 4,611 subjects, after dropping one subject for image corruption, for which PDFF values were calculated previously [20]. A PDFF value greater than 5.5% is the clinically accepted level for NAFLD. The generated ground truth data contained 919 subjects (19.9%) at a high risk for NAFLD. As alcohol consumption was a self-reported metric, we do not use it to filter subjects.

A second, larger UK Biobank cohort of 9,817 participants allowed us to include VAT and ASAT abdomen fat composition. In this cohort, the ground truth labels for PDFF, VAT, and ASAT were generated by [21]-[23]. The research described in this manuscript has been conducted using the UK Biobank Resource under application number 26041.

### B. Classification of NAFLD Risk

In this section, we describe our methodology to classify patients at a high-risk for NAFLD based on PDFF.

#### 1) Classification Model

For our initial exploration of the feasibility to train CNNs to classify high-risk NAFLD patients based on PDFF values, we performed an architecture search using popular 3D CNN variants. These four 3D CNN variants were selected based on state-of-the-art performance in similar biomedical imaging contexts. Using these architectures we further investigate the importance of channel interactions in diagnosing NAFLD.

##### ResNet

Our baseline implementation of 3D ResNet follows [24]. We include 3 layers of 3 residual blocks each, and extend the basic residual from [25] to 3D. A description of the layers and additional information relevant to building our network is described in Appendix A.

##### Non-local Neural Network

(referred as NL ResNet). As done in [26], we augment our baseline ResNet model using non-local operations whereby the response at a given position of a layer’s feature map is computed as the weighted sum of the features at all positions in the input feature maps. Non-local operations maintain the channel interactions of the baseline ResNet model and allow for capturing long-range dependencies along the *xyz* dimensions of the input volume.

##### ResNeXt

We alter our baseline ResNet model, by substituting the Basic ResNet blocks for Bottleneck blocks and introducing group convolutions in each 3 × 3 × 3 convolutional layers inside the Bottleneck block as done in [24, 27]. Group convolutions subset filters into groups and model each group independently, which sparsifies channel connections.

##### Channel-Separated Convolutional Network

(referred as CS ResNet). We extend the ResNeXt architecture as done in [28] by decomposing channel interactions from spatial interactions. We followed the (channel) interaction-preserved design of [28] by replacing the 3 × 3 × 3 Bottleneck layer convolution by a 1 × 1 × 1 convolutions for channel interactions followed by a *k* × *k* × *k* depth-wise convolutions for spatial interactions.

#### 2) Implementation Details

The dataset was split into train, validation, and test sets with 899, 100, and 3608 subjects respectively. Our models are randomly initialized and trained on single-channel volumes of size 160 × 160 × 10. We train for 50 epochs using early-stopping whenever validation loss does not improve for 4 epochs. We performed hyperparameter tuning on all models for number of layers, number of filters at each layer, strides, batch size, learning rate, and weight decay. Additionally, for NL ResNet, we experimented with the number and placement of non-local blocks, ultimately placing a single non-local block after the final residual layer. For ResNeXt, we experimented with the cardinality number, that is the number of groups in the 3 × 3 × 3 convolution inside the Bottleneck block and the ratio of conv2 layer input filters per group. For CS ResNet, we experimented with an interaction-preserved and interaction-reduced design as described in [28], the latter removing the 1 × 1 × 1 convolutions from the design described in the previous section. In the next section, optimal results of these scenarios are presented.

### C. Segmentation

We investigate the influence of the liver alone, as compared to the whole abdomen, towards classifying NAFLD. To achieve this, the liver was manually segmented by a non-expert from 50 subjects using MITK [29], and the annotated regions were visually validated by a clinician. The liver masks were split into train, validation, and test sets with 400, 50, and 50 slices respectively. The segmentation model was trained using a 2D U-Net deep neural network consisting of 23 convolutional layers as described in [12], and it was optimized using a weighted binary cross-entropy loss with β = 1.2 (Appendix B). Next, a post-processing filter which analyzes contiguous regions to account for inhomogeneities and a decision rule to quantify ratio of background to foreground pixels was applied.

Finally, this algorithm was extended to all the subjects in the dataset to create liver-only and sans-liver images for further analysis. This resulted in 4 subjects failing the quality check, and dataset size was reduced to 4,607 subjects. whenever validation loss does not improve for 4 epochs. To understand the determinants for model performance across the benchmarked architectures described in Section II B., we conduct additional experiments on NAFLD classification using liver-only and sans-liver as shown in Fig. 3.

### D. Abdomen Fat Composition

In the following part of the analysis, we fix the 3D CNN architecture to ResNet. We use the larger dataset containing VAT and ASAT values along with PDFF. We develop standalone and combination multitask regression models, and utilize the 5.5% threshold for PDFF to compute NAFLD classification metrics. As the values of each variable are non-negative and skewed (Fig. 1.),we fit all models in this part of the analysis using maximize likelihood based on a Gamma distribution and evaluate predictions across models and tasks using Spearman’s ρ.

**Fig. 1.**
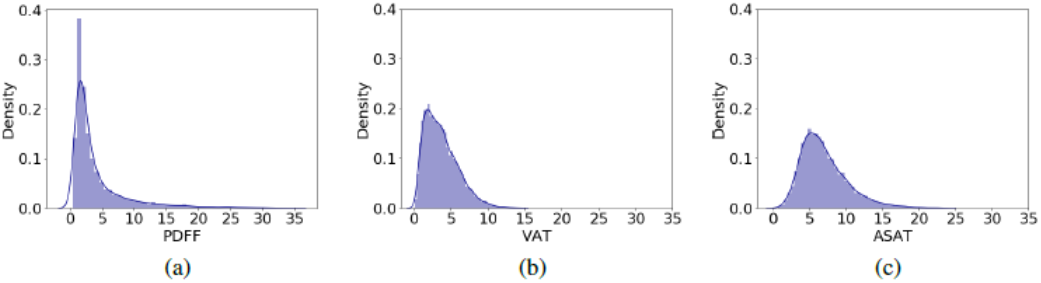
Dataset label distribution: (a) PDFF, (b) VAT, and (c) ASAT.

#### 1) Analyzing Task Relatedness

Initially, we developed separate regression models (Standalone) for each task, predicting PDFF, VAT, and ASAT raw values independently, to establish baseline performance. Next, we assessed the relationship between PDFF, VAT, and ASAT using pair-wise cosine similarities between the labels: 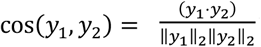 where *y*_1_ and *y*_2_ are PDFF, VAT, or ASAT label vectors.

Additional methods to assess task relatedness are explored in detail by Wu *et al*. in terms of task similarity, covariance, and model capacity [30]. To assess the task relationships empirically, we train three models on pairs of variables (PDFF-VAT, PDFF-ASAT, VAT-ASAT) and predict the raw values of the pairs of tasks. Additionally, we combine all three tasks, and train a single multitask model, predicting their values simultaneously. To minimize differences in the number of model parameters, we only change the final linear layer to output two or three predictors for the pair and multitask models, respectively. We refer to these models as multiple regression (MR). An MR model trained for PDFF and VAT is referred to as MR-PDFF-VAT. The combined model is represented in the results section as MR-ALL.

#### 2) Analyzing Multitask Learning Using Mixture-of-Experts

In the combined multitask model, MR-ALL, the intermediate representations are shared and only the final linear layer is specialized. We explore task similarity further by introducing expert layers and parameter sharing strategies. In particular, we implement a Multi-gate mixture-of-experts (MMoE) model as described by Ma *et al*. in [31]. Expert layers for the three tasks are added on top the shared ResNet from Section II B. Each expert block consists of three convolutional layers and a dropout. The features learned from each expert is shared in a fully connected manner using a softmax gate for every task as shown in Fig. 4(a). Intuitively, if the tasks are less related, then the softmax gates of the corresponding task would learn to utilize expert layers from the other tasks [31]. The softmax gates are simple linear transformation implemented using dense layers as described in [31] We investigate connecting the gates of experts across all task and only between the VAT and ASAT tasks, presented in Fig. 4(b). The reasoning behind this choice is explained in Section III.

#### 3) Implementation Details

The dataset was split into train, validation, and test sets with 4739, 500, and 4575 subjects respectively. Our models are randomly initialized and trained on single-channel volumes of size 160 × 160 × 10. We train for 30 epochs using the gamma loss described in equation (1) with *k* = 1.1 and *ε* = 1 × 10^−6^ and take the mean over the three labels.

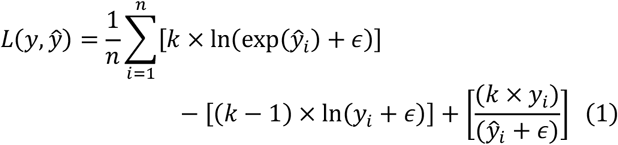

Where *y* ∈ ℝ^*n*^ and *ŷ* ∈ ℝ^*n*^ are the true and predicted label vectors for *n* samples, respectively. During training, the data set was bootstrapped and shuffled, and the average Spearman’s ρ values are presented in Table II and III.

## III. Results and Discussion

Our initial results training a ResNet on whole Abdomen MRI images to classify NAFLD using a PDFF value of > 5.5% demonstrated good performance, with Precision of 0.85 and F1 of 0.88 for NAFLD cases (see Table I). Examination of gradient activation maps from this condition demonstrated signal typically present in the liver for NAFLD patients (Fig. 5.).

**TABLE I.** NAFLD Classification Results

| Experiment | Model | NAFLD |  |  |
| --- | --- | --- | --- | --- |
|  |  | Precision | Recall | F1 |
| Abdomen | ResNet | 0.85 | 0.91 | 0.88 |
|  | **NL ResNet** | **0.88** | 0.91 | **0.89** |
|  | ResNeXt | 0.81 | 0.84 | 0.82 |
|  | CS ResNet | 0.78 | **0.92** | 0.85 |
| Liver-Only | ResNet | 0.67 | 0.56 | 0.61 |
|  | NL ResNet | 0.82 | 0.69 | 0.75 |
|  | ResNeXt | 0.64 | 0.57 | 0.60 |
|  | CS ResNet | 0.76 | 0.73 | 0.74 |
| Sans-Liver | ResNet | 0.55 | 0.27 | 0.36 |
|  | NL ResNet | 0.67 | 0.30 | 0.50 |
|  | ResNeXt | 0.42 | 0.61 | 0.50 |
|  | CS ResNet | 0.51 | 0.40 | 0.45 |

To understand the source of gradient activations in NAFLD subjects, we experimented with segmenting the liver from abdomen and examining NAFLD classification performance on liver only and sans-liver data. A 2D U-Net trained on the segmented data achieved a dice score of 0.93, which is comparable to results obtained in [13], [14] on segmentation tasks with dice score ranges from 0.85 to 0.94. Segmentation masks were applied to obtain liver-only and sans-liver images for every subject in addition to the whole abdomen MRI images, an example is shown in Fig. 2.

**Fig. 2.**
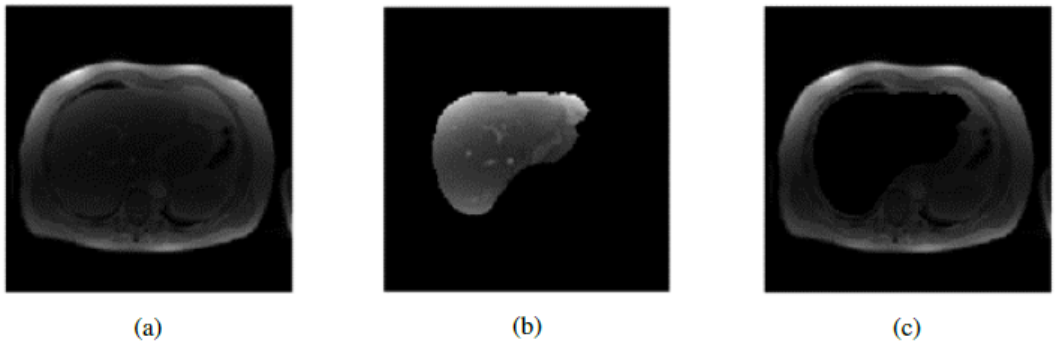
Example MRI slice of (a) Abdomen, (b) Liver-Only, and (c) Sans-Liver.

**Fig. 3.**
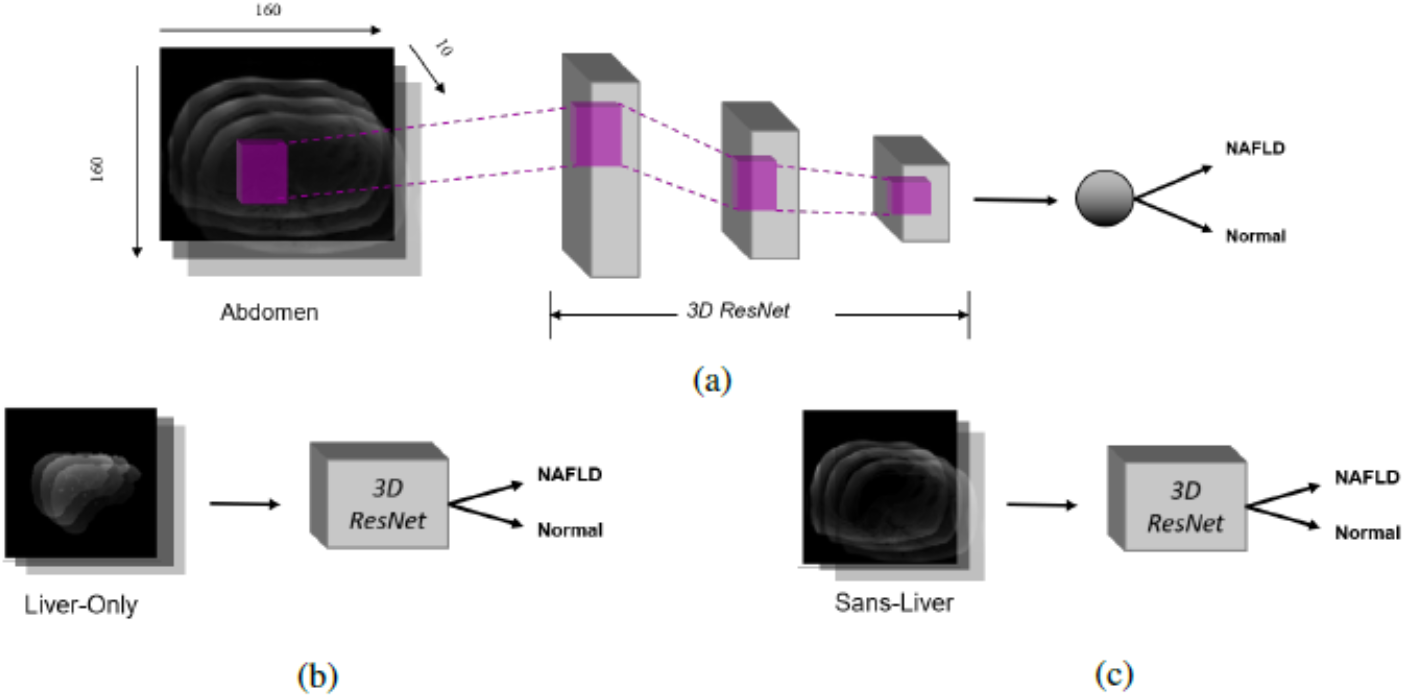
Workflow to classify NAFLD risk using the baseline ResNet from the three entities: (a)Abdomen, (b) Liver-Only, and (c) Sans-Liver.

**Fig. 4.**
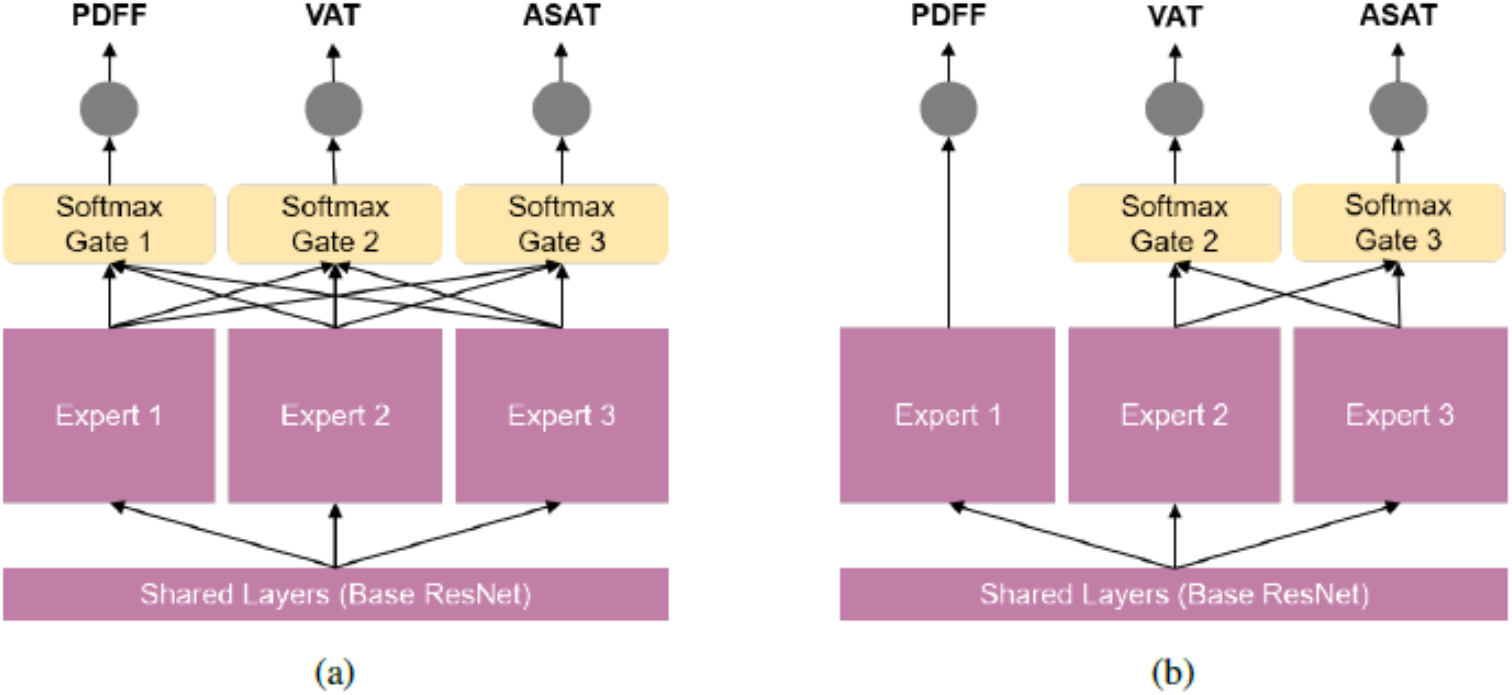
Estimating abdominal fat composition using: (a) Multi-gate Mixture-of-Experts (MMoE) and (b) Multi-gate Mixture-of-Experts-modified (MMoE-modified)

**Fig. 5.**
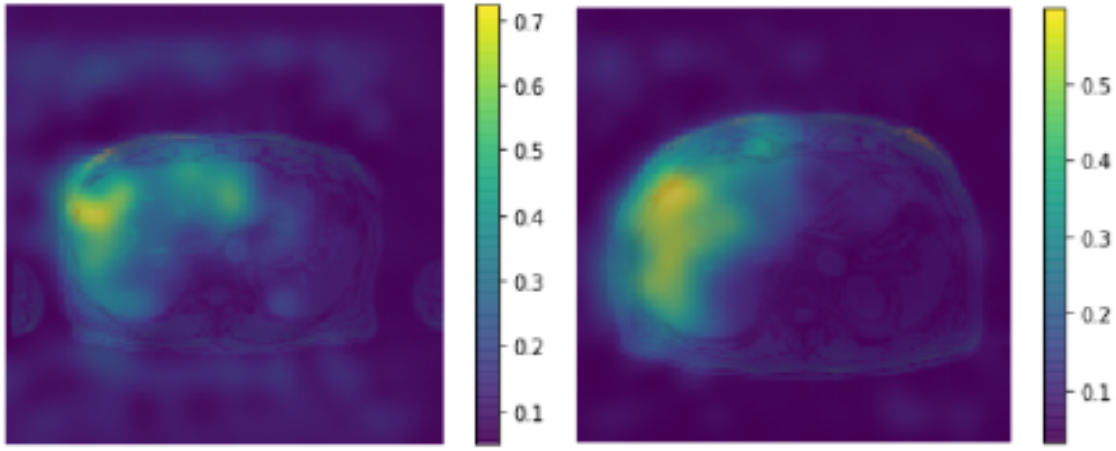
Examples of whole abdomen (slice) class activation maps for two subjects

The liver-only and sans-liver data were used to train the additional network architectures described in Section II B. to assess the influence of the liver in predicting NAFLD. We performed architecture search using four variants of the highly performant ResNet architectures on Abdomen, Liver-Only and Sans-Liver images. Interestingly, performance on whole abdomen MRI data demonstrated a higher Precision and F1 for NAFLD patients relative to segmented liver. NL ResNet was the highest performing architecture, demonstrating a Precision of 0.88 and F1 of 0.89 for NAFLD, suggesting the disease may manifest over long range dependencies across the *xyz* planes. Table I summarizes model performance across data and architectures.

In the second part of the analysis, we included VAT and ASAT measurements as well as PDFF, and used the larger dataset as described in section II D part 3. We fixed the model architecture to ResNet and fixed the optimization and training hyperparameters for all subsequent analyses. We evaluated each task independently (PDFF, VAT, ASAT) and the results are shown in the top of Table II. To evaluate task relatedness, we computed cosine similarity values between PDFF-VAT, PDFF-ASAT, and VAT-ASAT of 0.77, 0.72, and 0.87, respectively. This indicated high task similarity between VAT and ASAT.

**TABLE II.** Standalone and Multi-task Learning Results

| Model | $r_{PDFF}$ | $r_{VAT}$ | $r_{ASAT}$ |
| --- | --- | --- | --- |
| Standalone-PDFF | 0.91 | N/A | N/A |
| Standalone-VAT | N/A | 0.94 | N/A |
| Standalone-ASAT | N/A | N/A | 0.94 |
| MR-PDFF-VAT | 0.89 | 0.94 | N/A |
| MR-PDFF-ASAT | 0.89 | N/A | 0.93 |
| MR-VAT-ASAT | N/A | 0.94 | 0.93 |
| MR-ALL | 0.87 | 0.94 | 0.93 |
| MMoE | 0.91 | 0.94 | 0.94 |
| **MMoE-Modified** | **0.92** | **0.95** | 0.94 |

Note: Spearman's $\rho$ values for each variable with standard deviation values ranging from 0.00 to 0.07

Furthermore, these values were further supported by the change in Spearman’s ρ values between the standalone models and the pair models, noting a 0.02 decline in *r*_*PDFF*_ between standalone and pair models. This decline in PDFF prediction performance was further exacerbated in the multiple regression model between all three variables (MR-ALL), where Spearman’s ρ for PDFF declines by 0.04 against baseline standalone PDFF model.

Informed by this negative effect on PDFF, we studied the multitask Multi-gate Mixture-of-Experts(MMoE) architectures. We limited the negative effects and enabled positive knowledge transfer between the standalone tasks and the multitask MMoE by sharing experts and gates between the VAT and ASAT task and predicting PDFF with a separate network head in parallel, both on top of more robust shared intermediate representations. Results for the MMoE models are on the bottom of Table II.

To analyze whether the multitask models are learning distinct underlying representation or the correlation between the variables, we perform label permutation experiments and an ablation study using liver-only and sans-liver image data. To generate the larger segmentation dataset, the pre-trained segmentation model from section II C was applied to the 9,814 subjects. We first validated that the multitask results learn true shared representations and not task label correlations by permuting task labels. Intuitively, permuting labels breaks down the underlying correlation between tasks and, if the combined model was learning a label-correlation, then the overall performance would suffer when labels are permuted. As shown in Table V (Appendix C), performance for the permuted variable declines drastically, while that of the true (unpermuted) variables holds. We further study performance of the MMoE models on the liver-only and sans-liver data. Table III results show PDFF performance deterioration on the sans-liver data, similar to the effects in classification, however VAT and ASAT values remain unchanged suggesting MMoE models can robustly specialize the shared intermediate representation to each task.

**TABLE III.** Ablation Study: Estimating Abdominal Fat Composition

| Experiment | Model | $r_{PDFF}$ | $r_{VAT}$ | $r_{ASAT}$ |
| --- | --- | --- | --- | --- |
| Liver-Only | MMoE | 0.84 | 0.92 | 0.85 |
|  | MMoE-Modified | 0.85 | 0.92 | 0.86 |
| Sans-Liver | MMoE | 0.75 | 0.93 | 0.93 |
|  | MMoE-Modified | 0.76 | 0.94 | 0.94 |

Note: Spearman's $\rho$ values for each variable with standard deviation values ranging from 0.01 to 0.06

**TABLE IV.**
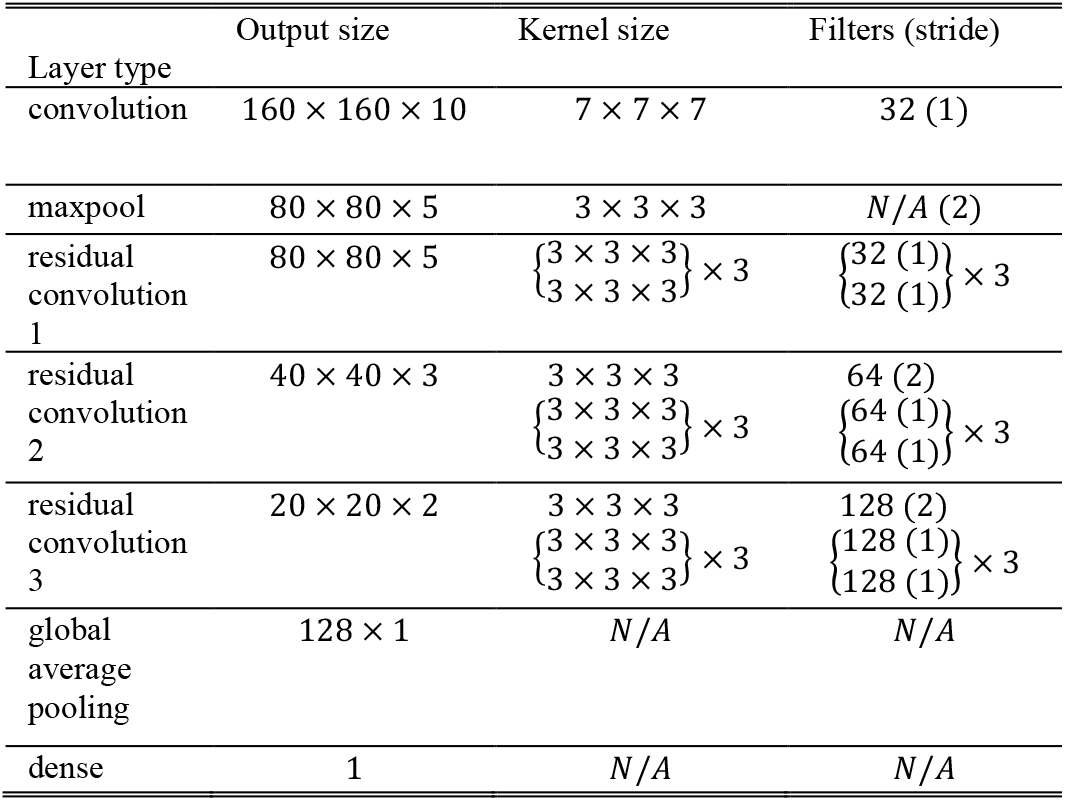
Baseline Resnet Architecture

**TABLE V.** Permutation Experiment Results to Analyze Feature Learning

| Model | Permuted label | $r_{PDFF}$ | $r_{VAT}$ | $r_{ASAT}$ |
| --- | --- | --- | --- | --- |
| MR-ALL | PDFF | 0.28 | 0.93 | 0.93 |
|  | VAT | 0.89 | 0.21 | 0.93 |
|  | ASAT | 0.89 | 0.94 | 0.04 |
| MMoE | PDFF | 0.36 | 0.94 | 0.94 |
|  | VAT | 0.91 | 0.22 | 0.93 |
|  | ASAT | 0.90 | 0.94 | 0.23 |
| MMoE-Modified | PDFF | 0.31 | 0.95 | 0.94 |
|  | VAT | 0.93 | 0.21 | 0.94 |
|  | ASAT | 0.92 | 0.95 | 0.30 |

## IV. Conclusion

Identifying patients at a high risk for NAFLD using a non-invasive MRI can provide significant benefits for patients, healthcare providers, and medical professionals, for example in early diagnosis, patient selection for clinical trials, and monitoring advanced forms of liver diseases in large populations. To the best of our knowledge, our work demonstrates the first machine learning classification model for NAFLD from MRI data applied to a large population in the UK Biobank dataset. Furthermore, applying a specialized multi-task learning model to enable positive task transfer between liver, visceral, and subcutaneous fat regression is applicable to other areas of medical imaging, where signal from several sources can be effectively disentangled and classified.

This research has several limitations. Classifying NAFLD using a PDFF threshold, while clinically accepted requires validation. The finding that models trained on the whole abdomen performed significantly better than segmented liver, despite class activation maps from whole abdomen demonstrating signal predominately in a patient’s liver, warrants further investigation. In future work, we plan to validate these findings in a larger cohort. We plan to explore genomic and clinical data from the UK Biobank in the context of NAFLD patients identified through this algorithm in order to clinical verify a distinct NAFLD radiomics phenotype.

## Data Availability

All data used is the manuscript is provided by the UK Biobank to registered researchers.

## Appendix

### A. Architecture of Baseline ResNet

### B. Loss Function

Weighted Binary Cross-Entropy:

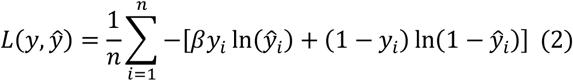

Where *y* ∈ ℝ^*n*^ and *ŷ* ∈ ℝ^*n*^ are the true and predicted label vectors for *n* samples, respectively. And β is set based on model tuning.

### C. Analyzing Label Correlation

## Acknowledgment

We thank our colleagues at AstraZeneca for fruitful discussions and especially Dr. Sudha Shankar, Dr. Faizal Khan, Dr. Ian Henry, and Dr. Claire Donoghue for their inputs on project planning and execution. Furthermore, we acknowledge the efforts of the open-source community to make tools available to the community and thereby foster science and transparency. Additionally, we thank members of the Scientific Computing Platform and Center for Genomics Research at AstraZeneca for providing the necessary resources for this project.

